# Targeted Muscle Reinnervation (TMR) or Regenerative Peripheral Nerve Interface (RPNI) for pain prevention in patients with limb amputation: A systematic review and meta-analysis

**DOI:** 10.64898/2026.11.15.26368167

**Authors:** David Gurpegui, Jesús del Moral, Montserrat Royo, Javier Pérez, Bernardo Hontanilla

## Abstract

**Background:** Phantom limb pain (PLP) and residual limb pain (RLP) after amputation impair rehabilitation, prosthetic use, and quality of life. Targeted Muscle Reinnervation (TMR) and Regenerative Peripheral Nerve Interface (RPNI) give transected nerves a physiologic target and may reduce chronic pain.

**Objective:** To synthesize evidence on prophylactic TMR and RPNI at amputation for preventing pain.

**Methods:** Following PRISMA 2020 and a published protocol (PROSPERO CRD42024617299), seven databases were searched to 31 July 2026 for studies of prophylactic TMR or RPNI at amputation reporting PLP, RLP, symptomatic neuroma, or safety outcomes. Pain intensity (Hedges’ g) and incidence (risk ratios, RR) were pooled separately for PLP and RLP using random-effects DerSimonian-Laird models. Risk of bias (ROBINS-I, RoB 2) and certainty (GRADE) were assessed.

**Results:** Twenty-seven studies (30 records, 2,154 participants; 16 comparative) were included. In comparative meta-analyses, TMR/RPNI was associated with lower pain intensity and incidence for PLP (g = -0.68, 95% CI -0.82 to -0.55; RR = 0.56, 0.48–0.65) and RLP (g = -0.68, -0.95 to -0.40; RR = 0.55, 0.42–0.73), or about 300 fewer patients with pain per 1,000 treated. Findings were directionally consistent and stable in sensitivity analyses. No study directly compared TMR with RPNI; indirect subgroup analyses showed no difference. Symptomatic neuroma was less frequent but imprecise (4 studies; RR = 0.35, 0.11–1.09); safety outcomes showed no clear differences. No study had low risk of bias; certainty was low to very low.

**Conclusions:** Prophylactic TMR and RPNI were associated with consistently less PLP and RLP than standard nerve management, remaining robust in sensitivity analyses excluding serious-risk-of-bias studies or weaker assumptions. Certainty remains low to very low due to observational designs, risk of bias, heterogeneous outcome definitions, and sparse secondary-outcome reporting. Prospective comparative studies with standardized outcomes are needed to compare techniques.

## Introduction

Limb amputation remains a major and growing source of long-term disability. Approximately 185,000 amputations are performed annually in the United States, and more than 3.6 million people are projected to be living with limb loss there by 2050 (1,2). Beyond the functional consequences of limb loss, post-amputation pain is among the most disabling sequelae. Persistent pain can interfere with rehabilitation, prosthetic use, sleep, mobility, work, and quality of life, and it is a common reason for prolonged opioid and neuropathic pain medication use (3,4). Post-amputation pain is commonly described in two overlapping but clinically distinct domains: phantom limb pain (PLP), perceived in the absent portion of the limb, and residual limb pain (RLP), localized in the stump of the remaining limb. Both are frequent: chronic PLP has been reported in more than 60% of patients, and symptomatic neuroma causing RLP in 4% to 49% of patients across recent systematic reviews (5,6).

The mechanisms underlying post-amputation pain are multifactorial. Peripheral nerve transection at the time of amputation may contribute particularly to RLP and may also provide ongoing peripheral input relevant to PLP. Central mechanisms, including spinal sensitization and cortical reorganization, can further maintain pain once established (7). Conventional nerve management during amputation, such as traction neurectomy or implantation into soft tissue, attempts to remove the nerve end from mechanical irritation but does not consistently provide a target for regenerating axons. As a result, traditional approaches may leave patients vulnerable to symptomatic neuroma, chronic pain, reduced prosthetic tolerance and subsequent surgeries (8–10).

Pharmacological management, physical therapy, and mirror therapy have yielded inconsistent results when used to prevent rather than to treat post-amputation pain, and once chronic neuropathic pain is established it responds poorly to any single modality (11,12). Attention has shifted toward modifying nerve handling at the amputation surgery, when the chance for neuroma formation occurs.

Targeted Muscle Reinnervation (TMR) and Regenerative Peripheral Nerve Interface (RPNI) were developed to address this problem by changing to active nerve management. TMR transfers transected sensory or mixed nerves to expendable motor branches of nearby muscles, allowing axons to reinnervate muscle targets. RPNI wraps the transected nerve end with an autologous free muscle graft, which could become available for reinnervation. Both techniques seek to give regenerating axons a target, reduce disorganized axonal sprouting, ectopic discharge, and symptomatic neuroma formation, and thereby decrease the peripheral input that can contribute to both PLP and RLP (13–15).

Initially used in selected patients for prosthetic control or for treatment of established neuroma pain, TMR and RPNI are increasingly performed at the time of primary amputation as prophylactic nerve management (16–19). Observational and comparative cohorts suggest that prophylactic active nerve management may reduce PLP, RLP, and symptomatic neuroma compared with standard amputation techniques (13). A recent multicenter analysis reported lower long-term opioid use and lower opioid dependence after primary TMR, without an increase in stump-related complications (20). However, the available evidence is heterogeneous. Studies vary in amputation level, etiology, timing of intervention, surgical technique, follow-up duration, pain instruments, outcome definitions, and whether pain is reported as incidence, severity, or categorical response.

Previous systematic reviews, including those by De Lange et al., Mauch et al., Yuan et al., and Zimbulis et al., support a protective effect of TMR and RPNI, but leave the prophylactic question incompletely answered (13,21–23). Yuan et al. examined primary TMR and RPNI performed at the time of amputation (23), and Zimbulis et al. compared TMR with standard nerve management (13). In these syntheses, however, PLP and RLP are frequently pooled into a single pain construct, reviews restricted to TMR cannot establish whether RPNI behaves similarly, and the evidence base has grown appreciably since their searches were completed. The present review differs in four respects. PLP and RLP are analyzed as separate domains. Pain incidence and pain intensity are treated as distinct estimands. Both TMR and RPNI are included, which allows the two techniques to be set against one another indirectly by subgroup, something reviews restricted to TMR cannot do. Finally, the certainty of the pooled evidence is rated using GRADE, and sensitivity analyses test how far the findings depend on risk of bias and on outcome-standardization assumptions.

This systematic review and meta-analysis aims to synthesize the available evidence on prophylactic TMR and RPNI performed at the time of limb amputation to prevent neuropathic pain. The primary objective is to compare TMR or RPNI with standard nerve management for PLP and RLP incidence and intensity. Secondary objectives are to summarize effects on symptomatic neuroma and safety outcomes, examine differences by intervention type, assess risk of bias and certainty of evidence, and identify limitations in the current evidence.

## Methods

### Protocol and Reporting

This systematic review and meta-analysis were conducted in accordance with the methodological guidance of the Cochrane Handbook for Systematic Reviews of Interventions and reported following the Preferred Reporting Items for Systematic Reviews and Meta-Analyses (PRISMA) 2020 statement (24–26). The protocol was prospectively registered in PROSPERO (CRD42024617299) and published before completion of the review (27).

### Eligibility Criteria

We included studies evaluating TMR or RPNI performed prophylactically at the time of limb amputation. Eligible populations comprised adults undergoing major upper- or lower-limb amputation for any indication. Studies of delayed or salvage TMR/RPNI for established post-amputation pain were excluded unless a prophylactic amputation-time arm could be extracted separately.

Eligible interventions were TMR or RPNI used as active peripheral nerve reconstruction at the time of amputation. Standard care was defined as conventional peripheral nerve management during amputation without TMR or RPNI, including traction neurectomy, ligation, implantation or burial of the nerve into soft tissue, muscle, or bone, myoplasty, or myodesis. We included randomized trials, non-randomized comparative studies, cohort studies, case-control studies, and case series with at least 8 participants. Conference abstracts and brief reports were eligible when they provided extractable clinical outcome data. Case reports, very small case series (n<8), narrative reviews, technical notes without extractable clinical outcomes, and studies not reporting relevant post-amputation pain or safety outcomes were excluded.

### Search Strategy and Study Selection

A comprehensive search was performed in electronic bibliographic databases from inception to 31 July 2026. Databases specified in the protocol included the Cochrane Central Register of Controlled Trials (CENTRAL), Web of Science, Scopus, PubMed/MEDLINE, CINAHL, Google Scholar, and MedRxiv, the latter two to identify unpublished/grey literature and minimize publication bias. Search terms combined concepts related to limb amputation, TMR, RPNI, peripheral nerve interface, nerve transfer, neuroma, phantom limb pain, residual limb pain, and post-amputation pain. No language restrictions were applied.

Records were screened independently by two reviewers (JdM, DG) using Covidence. Titles and abstracts were first assessed for potential eligibility, followed by full-text review of potentially relevant articles. Disagreements were resolved by discussion or consultation with a third reviewer. The study selection process was documented according to PRISMA and is shown in the PRISMA flow diagram.

**Figure 1.**
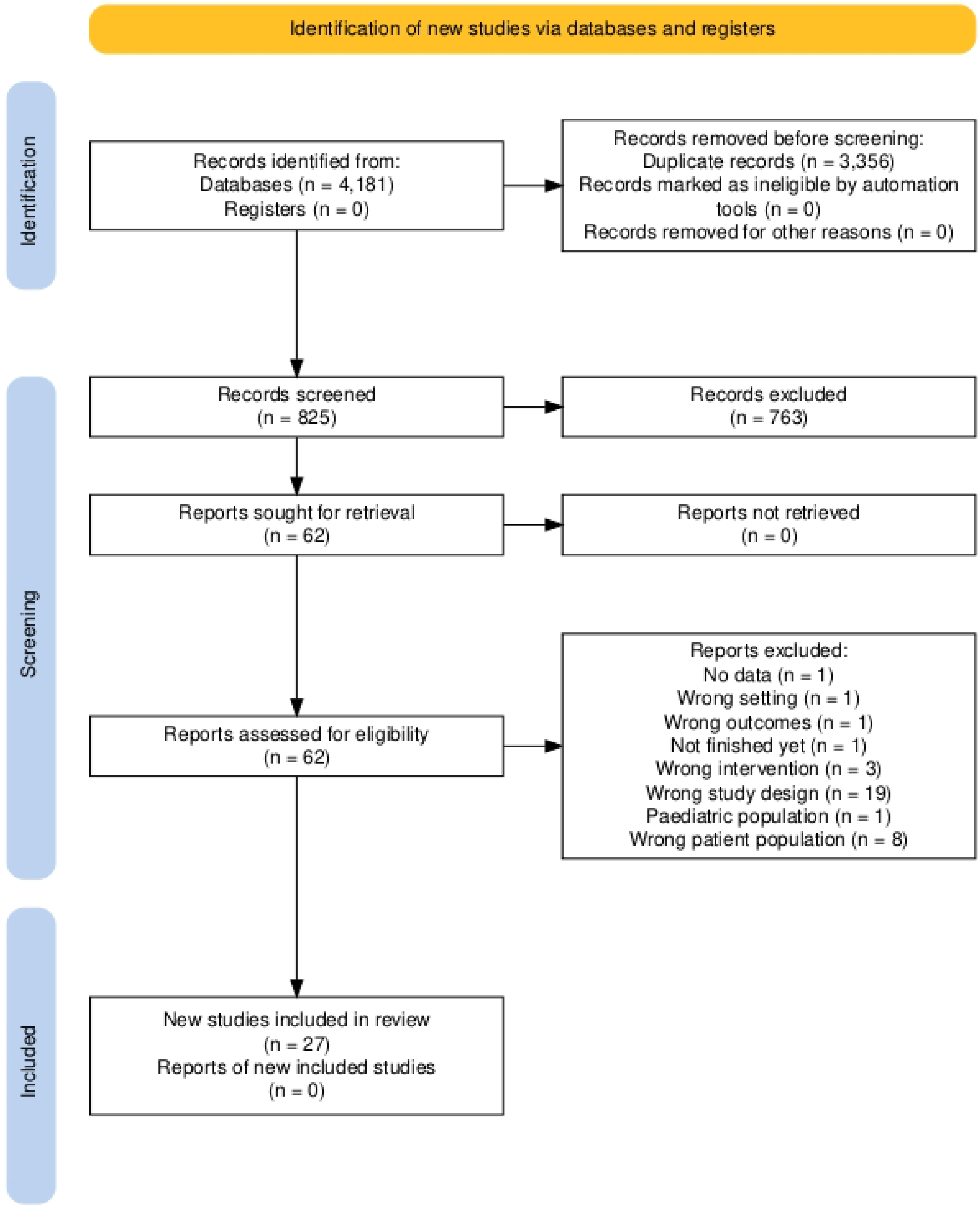
PRISMA 2020 flow diagram. Legend: Flow diagram showing identification, screening, eligibility assessment, and inclusion of studies. Searches were conducted from database inception to 31 July 2026.

### Data Extraction

Data were extracted using a structured data extraction form by three reviewers (DG, JdM, and MR). Each study was extracted independently by two of these reviewers, and disagreements were resolved by consensus or by consultation with the third. Extracted study characteristics included author, year of publication, study design, sample size, follow-up duration, amputation level, amputation etiology, intervention type, surgical timing, and comparator details. Patient-level aggregate characteristics included age, sex, comorbidities, smoking status, body mass index, vascular disease, diabetes, and other clinically relevant baseline factors when reported.

Outcome data were extracted separately for TMR/RPNI intervention groups and standard care controls. Primary efficacy outcomes were phantom limb pain (PLP) and residual limb pain (RLP), each extracted as both pain intensity and a binary pain outcome whenever possible. Because the clinical question was prophylaxis, comparative binary analyses were reported as incidence analyses; single-arm summaries were reported as prevalence or pain burden.

Secondary outcomes included symptomatic neuroma, hematoma, infection, wound dehiscence, cardiovascular events, reoperation, and mortality. When required data were reported only in figures, values were extracted from graphical displays and retained with source notes. When several pain instruments or time points were available, standardized extraction rules were used to select the outcome most representative of the relevant post-amputation pain domain.

### Risk of Bias Assessment

Risk of bias was assessed according to study design, independently and in duplicate by the same three reviewers, with disagreements resolved by consensus. Randomized trials were assessed using the Cochrane Risk of Bias 2 tool (28) and non-randomized comparative studies using ROBINS-I (29). Study records without an eligible standard-care control group were not assigned a formal risk-of-bias judgement and were labelled as not assessed because the planned comparative risk-of-bias framework was not applicable. Domain-level judgements were recorded, together with an overall risk-of-bias judgement categorized as low, moderate, or serious when sufficient information was available. Risk-of-bias judgements were summarized descriptively and used in sensitivity analyses excluding studies judged to be at serious overall risk of bias.

### Outcome Standardization and Assumptions

Pain outcomes required standardization because included studies used heterogeneous instruments, time points, and reporting formats. PLP and RLP were analyzed separately. For each pain domain, two outcome types were created: continuous pain intensity and a binary pain outcome.

Pain intensity was standardized to a 0-10 metric. Visual Analog Scale (VAS) or Numeric Rating Scale (NRS) scores reported on a 0-10 scale were used directly. PROMIS T-scores were transformed to an approximate 0-10 scale using the formula (T-score - 30)/4. When multiple PROMIS domains were available, PROMIS Pain Intensity was preferred, followed by Pain Interference and Pain Behavior. When studies reported pain using ordinal categories only, category midpoints were used to estimate a weighted mean: no pain = 0, mild pain = 2, moderate pain = 5, and severe pain = 8.5.

Means and standard deviations were extracted directly whenever available. Medians were used as approximations of means when means were not reported. Standard deviations were extracted directly when available; when only interquartile ranges were reported, standard deviations were approximated as IǪR/1.35. For PROMIS-derived outcomes, standard deviations were rescaled using the same linear transformation as the mean. In selected studies where only binary pain incidence was available but a continuous synthesis required an approximate pain-intensity value, a conservative binary-to-mean transformation was used, assigning a score of 5 to patients with pain and 0 to patients without pain; the corresponding standard deviation was derived from the binomial distribution on this 0-5 contrast. These transformations were recorded at study level and evaluated in sensitivity analyses.

In a minority of studies, the standard deviation used for the standardized pain score, the no-pain proportion or both were not directly reported by the source publication and were carried forward from the structured extraction as imputed values. This applied to Alexander 2019, Frantz 2020, Kumar 2024, Raasveld 2024, Reid 2024, Smith 2024, Elmorsi 2025, and Mathieu 2025 (30–37). Every imputed value was flagged in the study-level source field of the analytic dataset so that its origin remains traceable, and the influence of these assumptions was examined in the sensitivity analyses described below.

The binary pain outcome was expressed as the proportion of patients with pain versus no pain when this was directly reported. When a strict no-pain definition was unavailable but clinically meaningful categories were reported, study-specific transformations were applied and documented. For example, in studies reporting categorical pain severity, no pain or no clinically significant pain was used when this best matched the published data structure. Where a study reported a mean pain score but no usable no-pain category, the proportion was reverse-imputed as one minus the mean divided by five, the inverse of the binary-to-mean rule above; this applied only to Smith 2024 (35). All such assumptions were tracked in the dataset.

### Statistical Analysis

All analyses were performed in Stata version 16 (StataCorp, TX, USA) (38). Comparative meta-analyses were conducted when at least two studies contributed analyzable data for a given outcome. Continuous outcomes were pooled as standardized mean differences using Hedges’ g with 95% confidence intervals, as anticipated when studies used different pain instruments. Binary outcomes were pooled as risk ratios rather than odds ratios: because phantom and residual limb pain are common events, odds ratios diverge from risk ratios, overstate the apparent magnitude of effect, and are frequently misinterpreted as risk ratios. Risk ratios were therefore considered the more appropriate and clinically interpretable measure, and a continuity correction for zero cells was applied as implemented by Stata’s meta-analysis commands. Random-effects models using the DerSimonian-Laird method were used throughout because clinical and methodological heterogeneity was expected across studies (39).

Some studies contributed more than one analytic record, either because technique-specific cohorts were reported separately or because different outcomes were reported in different populations. Records from the same study were never entered into the same comparative meta-analysis. In the supplementary single-arm syntheses, two studies contributed two records each (Obinero 2025 and Mahmoud 2024) (40,41); in both cases the records correspond to disjoint groups of patients, so no participant contributed more than once to any pooled estimate. Where two records from the same study described overlapping populations, participants were counted once in the descriptive totals: the 12-month PLP record of Berberoglu 2025 (47 patients) is a nested subset of its neuroma record (62 patients), and only the larger cohort contributes to the reported participant counts (42).

The main comparative analyses evaluated TMR or RPNI versus standard care for PLP intensity, PLP incidence, RLP intensity, and RLP incidence. Forest plots were generated for all primary meta-analyses. Statistical heterogeneity was assessed using the Ǫ test, tau-squared, and I-squared. I-squared values were described using the approximate bands suggested in the Cochrane Handbook (24), in which 0% to 40% may not be important, 30% to 60% may represent moderate heterogeneity, 50% to 90% substantial heterogeneity, and 75% to 100% considerable heterogeneity. These ranges overlap by design and were read together with tau-squared and the Ǫ test rather than applied as thresholds. Funnel plots and Egger regression tests for small-study effects were generated for the four main comparative pain outcomes, the Egger test being implemented as a regression of the standard normal deviate on precision with a test of the intercept. These tests were interpreted cautiously because each analysis included 11 studies and the evidence was clinically heterogeneous.

No included study compared TMR with RPNI directly. Because each technique was nonetheless compared against standard care, the two were compared indirectly: each of the four main analyses was repeated as a subgroup meta-analysis by intervention type, with a chi-square test for subgroup differences, and the subgroups are displayed in the corresponding forest plots. Such comparisons are observational at the study level, since the studies contributing each technique differ in population, setting, and era, and few studies evaluated RPNI. They were therefore planned and interpreted as exploratory rather than as a test of one technique against the other.

### Secondary and Supplementary Analyses

Single-arm pooled analyses were performed for intervention groups and control groups separately when comparative synthesis was not possible or when summarizing the broader evidence base was informative. For single-arm prevalence analyses, study-level proportions were analyzed with a small continuity adjustment to stabilize estimates in the presence of very high or very low event rates. Single-arm mean pain scores were pooled using study-level standard errors derived from the reported or imputed standard deviations.

Secondary outcomes were synthesized when at least two comparative studies reported the outcome. These comprised symptomatic neuroma and the safety outcomes hematoma, infection, wound dehiscence, cardiovascular events, reoperation, and mortality. All were analyzed as risk ratios using random-effects models.

### Sensitivity Analyses

Sensitivity analyses were conducted to evaluate the robustness of the findings. First, analyses were repeated after excluding the weakest imputations. For pain intensity, this removed the binary-to-mean transformation applied to studies reporting only pain incidence (Kubiak 2019, Chang 2021, and Chang 2024) (43–45), the incidence-by-severity population-mean approximation (Chang 2024) (45), and acute postoperative VAS values not specific to chronic PLP or RLP (Smith 2024) (35). For pain incidence, this removed studies whose incidence-like endpoint was not a strict no-pain definition, namely those using no or mild pain (Berberoglu 2025) or absence of severe pain (Pejkova 2022) as the no-pain category (42,46), those whose no-pain proportion was reverse-imputed from a mean score (Smith 2024) (35), and those contributing pain scores only (Mathieu 2025) (37). Second, analyses were repeated excluding studies rated as serious overall risk of bias when risk-of-bias data were available.

### Certainty of Evidence

Certainty of evidence for the main outcomes was assessed using the GRADE framework by the same three reviewers (DG, JdM and MR) (47), with the final rating for each outcome agreed by consensus. Because most included studies were observational or non-randomized, certainty ratings started at low certainty unless randomized evidence made a higher starting point appropriate. Downgrading considered risk of bias, inconsistency, indirectness, imprecision, and potential publication bias. Outcome-scale standardization itself was not treated as a primary reason for downgrading when sensitivity analyses excluding weaker imputations and less directly comparable outcome sources yielded similar conclusions.

## Results

### Study Selection

The electronic search identified 4,181 records across databases. After removal of 3,356 duplicate records, 825 records were screened by title and abstract, of which 763 were excluded. Sixty-two reports were retrieved and assessed for eligibility. Thirty-five full-text reports were excluded, most commonly because of wrong study design (n = 19), wrong patient population (n = 8), wrong intervention (n = 3), no extractable data (n = 1), wrong setting (n = 1), wrong outcomes (n = 1), pediatric population (n = 1), or an unfinished study (n = 1). Twenty-seven studies were included in the review (30–37,40–46,48–59). The final quantitative dataset included 30 study records published between 2014 and 2025, comprising 2,154 unique participants. Of these, 985 participants underwent TMR or RPNI and 1,169 were included in standard care or comparator groups.

### Study Characteristics

Seventeen study records, corresponding to 16 studies, included a comparative standard care group and contributed 1,685 unique participants to comparative analyses. The remaining 13 records were single-arm intervention cohorts and were retained for descriptive and supplementary single-arm syntheses when relevant outcome data were available. Most study records evaluated TMR (23 records), while 7 evaluated RPNI.

Of the 27 included studies, 21 were retrospective, 5 were prospective or used prospectively collected registry data, and 1 was an open randomized controlled trial. Four studies were available only as conference abstracts (Kumar 2024, Mahmoud 2024, Elmorsi 2025, and Vaeth 2025) (32,36,41,51). Study size varied widely, with a median of 56 participants per analytic record (range 3 to 483). Reported mean age in intervention groups ranged from 34 to 61 years, and the proportion of male participants ranged from 42% to 83%. Pain was measured with the Numeric Rating Scale, the Visual Analog Scale, PROMIS instruments, or ordinal severity categories, and the timing of outcome assessment ranged from 6 weeks to 24 months after amputation. Included studies also varied in amputation level, surgical indication, and intervention timing.

**Table 1.** Characteristics of included comparative studies.

| Study (Author, Year) | Surgery | Design | Sample Size (Interv / Ctrl) | Follow-up | Outcomes | Risk of Bias |
|---|---|---|---|---|---|---|
| Alexander 2019 (30) | TMR | Retrospective cohort | 85 (27 / 58) | > 1 yr | Symptomatic neuroma; PROMIS (Intensity, Behavior, Interference) for PLP/RLP; | Serious |
| Kubiak 2019 (43) | RPNI | Retrospective matched cohort | 90 (45 / 45) | mean ~357 d | Symptomatic neuroma and PLP; no continuous scale | Serious |
| Valerio 2019 (53) | TMR | Cohort with IPTW-adjusted external control | 483 (43 / 440) | median follow-up 330 d | NRS for PLP/RLP; PROMIS (Intensity, Behavior, Interference) | Moderate |
| Chang 2021 (44) | TMR | Cohort with historical control | 200 (100 / 100) | mean 9.6 mo (TMR) vs. 18.5 mo (Ctrl) | Presence of RLP and PLP; NRS pain severity; complications | Moderate |
| O'Brien 2021 (48) | TMR | Retrospective cohort with benchmark control | 71 (16 / 55) | mean follow-up 23.1 mo (range 6–65) | NRS for PLP/RLP; PROMIS (Intensity, Behavior, Interference) | Moderate |
| Huffman 2022 (49) | TMR | Retrospective cohort | 51 (8 / 43) | 30-d complications, 1-yr mortality; pain at 6 mo | Complications; NRS nonspecific pain | Moderate |
| Pejkova 2022 (46) | RPNI | Retrospective comparative case series | 28 (14 / 14) | mean 49 wk (range 24–76) | PROMIS (Interference); VAS severity categories at 24 wk | Serious |
| Shammas 2022 (50) | TMR | Retrospective propensity score-matched cohort | 96 (31 / 65) | 60-d postoperative complications | Complications | Moderate |
| Lu 2023 (52) | RPNI-like (NI) | Retrospective cohort | 44 (8 / 36) | Questionnaire at 12 mo; mean 74 wk (NI) vs. 107 wk (Ctrl) | NRS for PLP/RLP; PROMIS (Intensity, Behavior, Interference) | Moderate |
| Chang 2024 (45) | TMR | Retrospective cohort | 99 (41 / 58) | Mean 9.5 mo (TMR) vs. 14.3 mo (Ctrl) | NRS for RLP/PLP (severity); complications | Moderate |
| Kumar 2024* (32) | TMR | Open RCT | 40 (20 / 20) | At 12 wk | NRS for RLP/PLP | Serious (RoB 2) |
| Smith 2024 (35) | TMR | Retrospective cohort | 103 (28 / 75) | Neuroma: median 1.2 yr (0–7.2); VAS at 6 wk | Symptomatic neuroma; VAS 0–10 at 6 wk (not specific) | Serious |
| Berberoglu 2025a† (42) | RPNI | Retrospective cohort | 62 (27 / 35) | Mean 26.3 mo (range 3–58.5) | Symptomatic neuroma | Serious |
| Berberoglu 2025b† (42) | RPNI | Retrospective cohort | 47 (23 / 24) | PLP assessed at 12 mo | NRS PLP categorized as no/mild (0–3), moderate (4–6), severe (7–10) | Serious |
| Elmorsi 2025* (36) | TMR RPNI§ | Retrospective cohort | 88 (55 / 33) | 381 d (TMR/RPNI) vs. 1,743 d (Ctrl) | NRS for PLP/RLP; PROMIS (Intensity, Behavior, Interference) | Serious |
| Mathieu 2025 (37) | TMR | Prospective cohort | 28 (10 / 18) | Median 14.5 mo (IQR 13–21, TMR) vs. 26 mo (IQR 10.5–46.5, Ctrl) | NRS for PLP/RLP; PROMIS (Intensity, Behavior, Interference) | Moderate |
| Vaeth 2025* (51) | TMR | Retrospective cohort | 117 (43 / 74) | 12 mo required; median time to neuroma: 717 d (TMR) vs. 573 d (Ctrl) | Symptomatic neuroma | Serious |

**Footnotes for Table 1:**
\* Conference abstract.
**†** In Berberoglu 2025 (42), record *a* (*n* = 62) represents the symptomatic neuroma cohort and record *b* (*n* = 47) represents a nested subset with available 12-month PLP data; only the larger cohort (*n* = 62) is counted toward the total unique participants.
**§** Elmorsi 2025 (36) reported a combined TMR/RPNI cohort without a technique-specific split and was coded under TMR for mutually exclusive subgroup fields.

Outcome availability varied across pain domains and analysis types. For PLP, 19 intervention cohorts reported analyzable pain intensity data and 18 reported analyzable binary pain data. Comparative PLP analyses included 11 studies for intensity and 11 studies for incidence. For RLP, 21 intervention cohorts reported analyzable pain intensity data and 19 reported analyzable binary pain data. Comparative RLP analyses included 11 studies for intensity and 11 studies for incidence.

### Risk of Bias

Across the 27 included studies, overall risk of bias was classified as moderate in 8 studies (29.6%), serious in 8 studies (29.6%), and not assessed in 11 studies (40.7%) because no eligible standard-care control group was extracted. No study was classified as low overall risk of bias.

Among the 16 studies with a formal assessment, confounding was the dominant limitation: it was rated serious in 7 studies and moderate in 8, and only 1 study was judged to be at low risk in this domain. This reflects the predominance of non-randomized designs in which allocation to nerve reconstruction was determined by surgeon preference, institutional practice, or calendar period. Measurement of outcomes was rated moderate in 13 of the 16 studies, mainly because outcome ascertainment was retrospective and unblinded. By contrast, selection of participants, classification of interventions, and deviations from intended interventions were rated low in most studies. Risk-of-bias summaries by domain and overall judgement are provided in the supplementary figures and tables.

### Primary Comparative Outcomes

TMR or RPNI was associated with lower PLP intensity compared with standard care. Eleven comparative studies contributed to this analysis. The pooled standardized mean difference was Hedges’ g = -0.68 (95% CI -0.82 to -0.55; p < 0.001), favoring TMR/RPNI. There was no between-study heterogeneity (I2 = 0.0%; tau2 = 0.0000).

**Figure 2.**
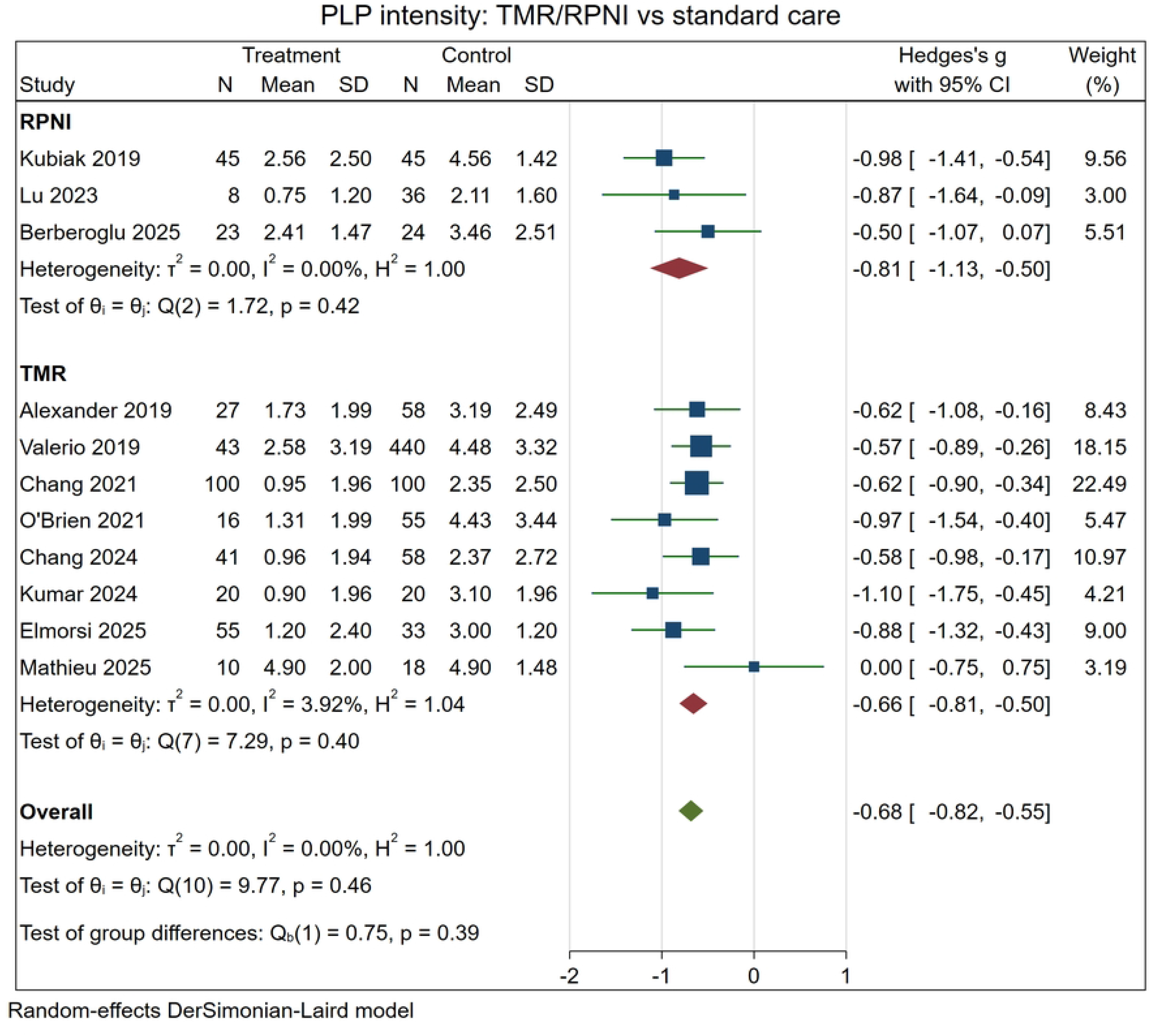
Phantom limb pain intensity: TMR/RPNI versus standard care. Legend: Forest plot of comparative studies reporting PLP intensity. Effect sizes are Hedges’ g with 95% CI. Negative values favor TMR/RPNI. The pooled random-effects estimate favored TMR/RPNI (11 studies; Hedges’ g = -0.68, 95% CI -0.82 to -0.55; I2 = 0.0%).

TMR or RPNI was also associated with lower PLP incidence. Eleven comparative studies contributed to the PLP incidence analysis. The pooled risk ratio was 0.56 (95% CI 0.48 to 0.65; p < 0.001), indicating a lower risk of PLP among patients treated with TMR or RPNI compared with standard care. Heterogeneity was negligible (I2 = 0.0%; tau2 = 0.0000).

**Figure 3.**
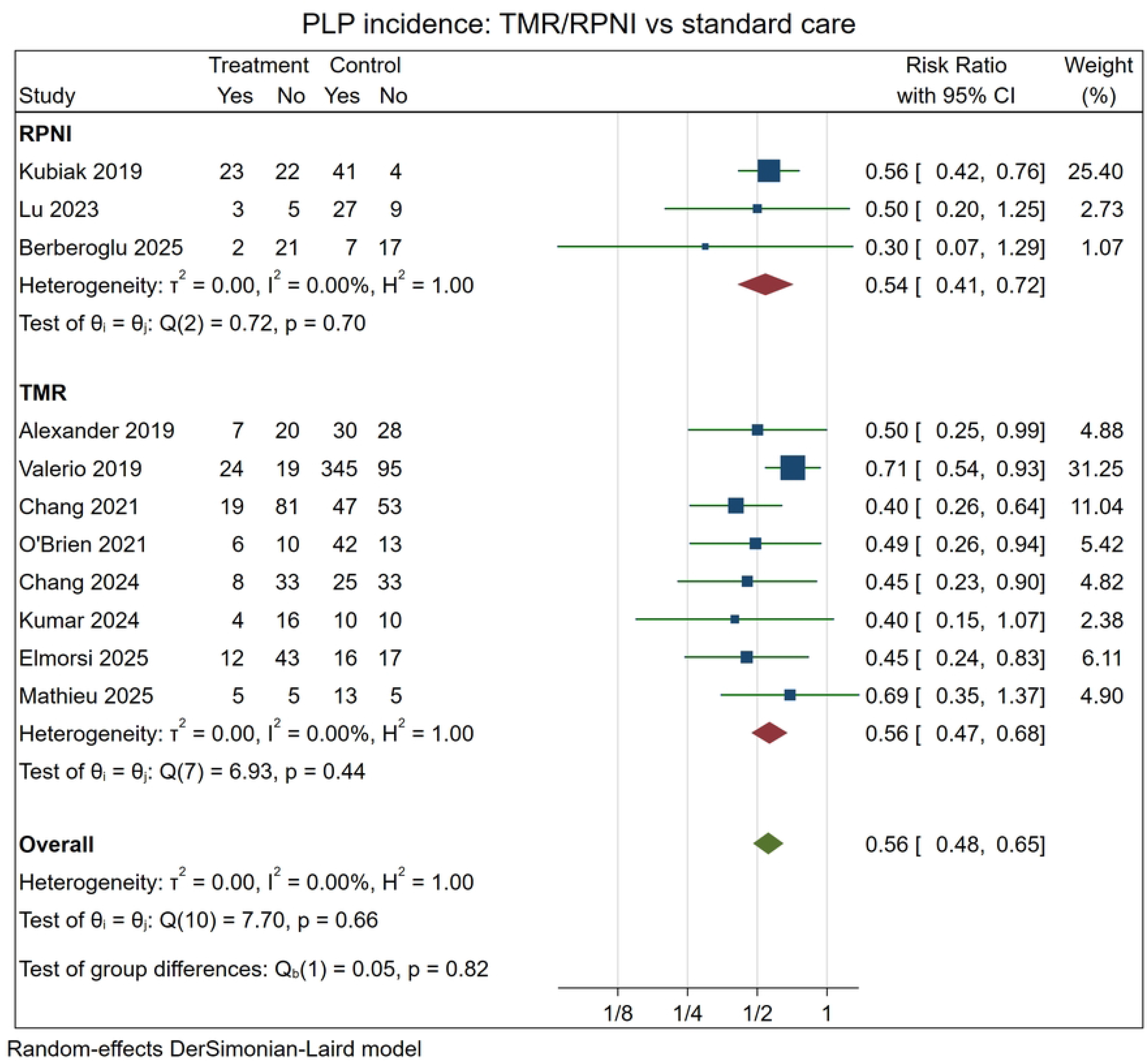
Phantom limb pain incidence: TMR/RPNI versus standard care. Legend: Forest plot of comparative studies reporting PLP incidence. Effects are risk ratios with 95% CI. Values below 1 favor TMR/RPNI. The pooled random-effects estimate favored TMR/RPNI (11 studies; RR = 0.56, 95% CI 0.48 to 0.65; I2 = 0.0%).

For RLP intensity, 11 comparative studies were included. TMR or RPNI was associated with lower RLP intensity compared with standard care, with a pooled Hedges’ g of -0.68 (95% CI -0.95 to -0.40; p < 0.001). Heterogeneity was substantial (I2 = 71.3%; tau2 = 0.1399), reflecting clinical and methodological variability across studies and outcome definitions. One included study (Smith 2024) reported slightly higher pain in the TMR group than in controls; this estimate came from acute overall postoperative VAS at six weeks rather than from RLP-specific chronic pain.

**Figure 4.**
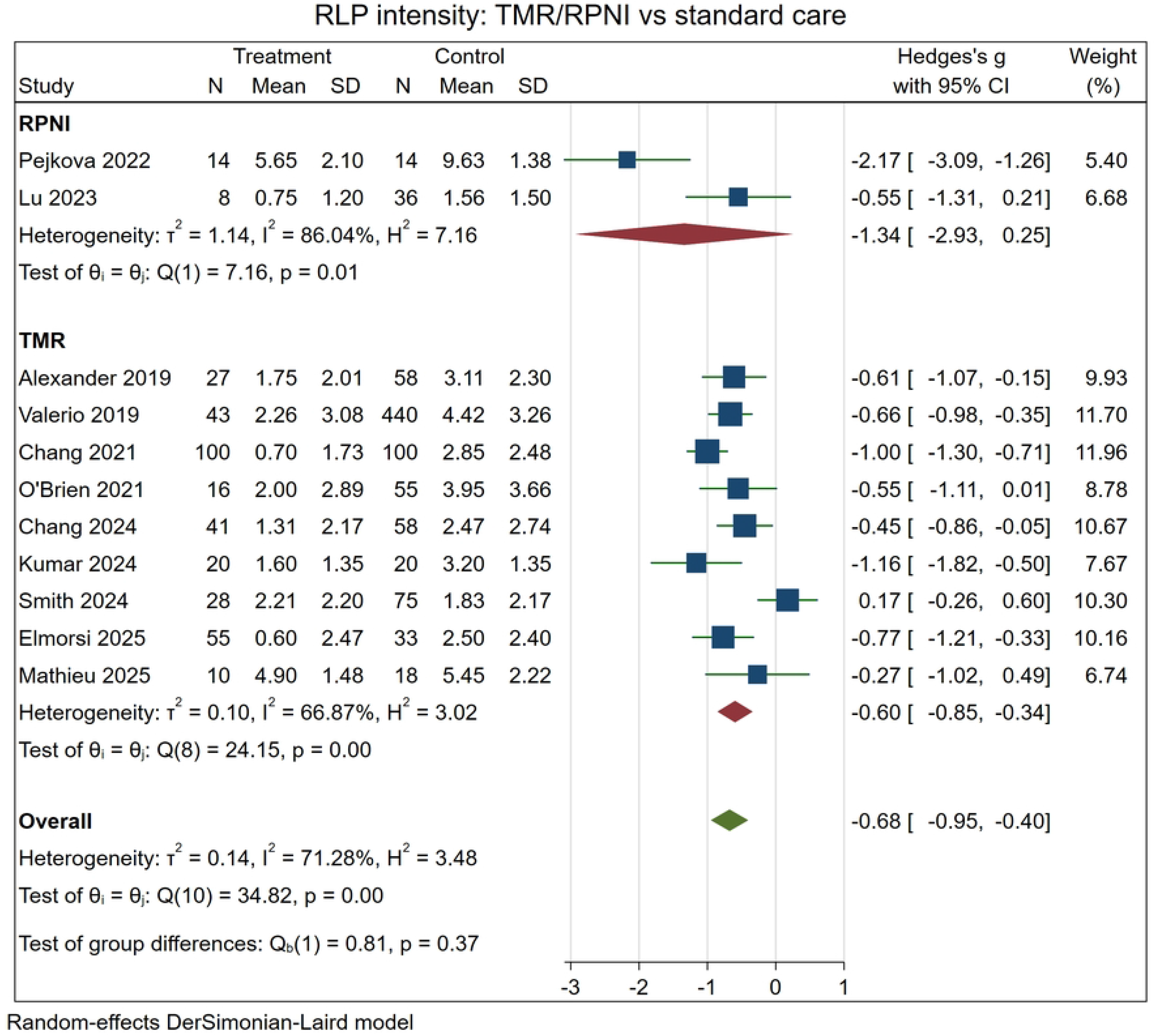
Residual limb pain intensity: TMR/RPNI versus standard care. Legend: Forest plot of comparative studies reporting RLP intensity. Effect sizes are Hedges’ g with 95% CI. Negative values favor TMR/RPNI. The pooled random-effects estimate favored TMR/RPNI (11 studies; Hedges’ g = -0.68, 95% CI -0.95 to -0.40; I2 = 71.3%).

For RLP incidence, 11 comparative studies contributed data. TMR or RPNI was associated with lower RLP incidence compared with standard care, with a pooled risk ratio of 0.55 (95% CI 0.42 to 0.73; p < 0.001). Heterogeneity was moderate to substantial (I2 = 59.0%; tau2 = 0.1178).

**Figure 5.**
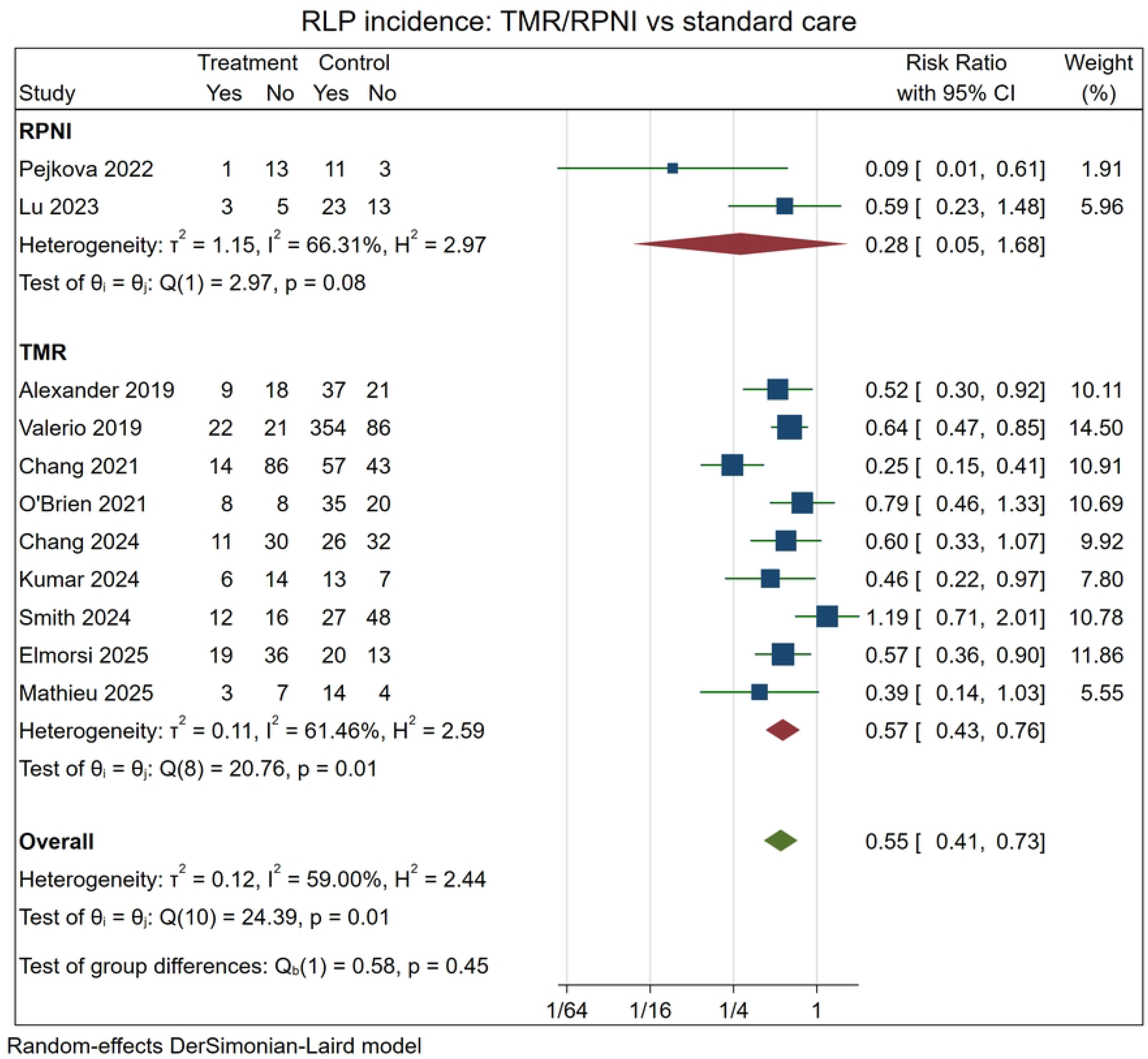
Residual limb pain incidence: TMR/RPNI versus standard care. Legend: Forest plot of comparative studies reporting RLP incidence. Effects are risk ratios with 95% CI. Values below 1 favor TMR/RPNI. The pooled random-effects estimate favored TMR/RPNI (11 studies; RR = 0.55, 95% CI 0.42 to 0.73; I2 = 59.0%).

The PLP analyses included 1,275 participants and the RLP analyses 1,269 participants. Applied to the observed control risk of 680 per 1000, which was the same in both pain domains, the pooled risk ratios correspond to 379 per 1000 for PLP (95% CI 326 to 441), or 301 fewer per 1000 (95% CI 239 to 354 fewer), and to 374 per 1000 for RLP (95% CI 286 to 496), or 306 fewer per 1000 (95% CI 184 to 394 fewer). Expressed as an approximate number needed to treat for benefit, these absolute effects correspond to about 4 patients treated to prevent one patient with PLP and about 4 patients treated to prevent one patient with RLP, assuming the observed control-group risk applies (3.32 and 3.27 before rounding up).

The direction of effect was consistent at study level. Of the 44 study-level estimates contributing to the four primary analyses, 41 favored TMR or RPNI, one was exactly neutral (Mathieu 2025, PLP intensity) (37), and the only two favoring standard care both came from Smith 2024 (35). All 11 studies contributing to the PLP incidence analysis favored TMR or RPNI.

### Comparison by Intervention Type

No included study compared TMR directly with RPNI. Because both techniques were compared against standard care, however, the two technique-specific estimates can be placed side by side as an indirect comparison, and these subgroups are displayed in the main forest plots. The PLP analyses comprised 8 TMR and 3 RPNI studies, and the RLP analyses 9 TMR and 2 RPNI studies. For PLP incidence the two estimates were almost superimposable (TMR: RR 0.56, 95% CI 0.47 to 0.68; RPNI: RR 0.54, 95% CI 0.41 to 0.72; test for subgroup difference p = 0.82), with no heterogeneity within either subgroup. For RLP incidence the RPNI estimate rested on two studies and was correspondingly imprecise (TMR: RR 0.57, 95% CI 0.43 to 0.76; RPNI: RR 0.28, 95% CI 0.05 to 1.68; test for subgroup difference p = 0.45).

Pain intensity showed the same pattern. For PLP, the pooled Hedges’ g was -0.66 (95% CI -0.81 to -0.51) with TMR and -0.81 (95% CI -1.13 to -0.50) with RPNI (test for subgroup difference p = 0.39); for RLP, it was -0.60 (95% CI -0.85 to -0.34) with TMR and -1.34 (95% CI -2.93 to 0.26) with RPNI (p = 0.37). Confidence intervals overlapped in every comparison, and no test for subgroup differences approached significance.

These comparisons should be read as indirect and exploratory. They are observational at the study level, because the TMR and RPNI studies differ in population, setting, and era; the RPNI subgroups contain only two to three studies; and one of the two RPNI studies contributing to RLP (Pejkova 2022) is the principal source of heterogeneity in that analysis (46). The data therefore provide no evidence that the two techniques differ in effect—particularly for PLP, where the subgroup estimates were nearly identical—although they do not establish equivalence

### Single-Arm and Supplementary Pain Analyses

Single-arm syntheses were conducted to summarize pain burden in intervention and control cohorts separately when sufficient data were available. These analyses are reported as supplementary because they do not preserve within-study comparisons and are more vulnerable to differences in case mix, follow-up, and outcome measurement.

Pooled PLP prevalence was 30.3% (95% CI 23.5% to 37.1%) across 18 intervention cohorts and 60.8% (95% CI 49.6% to 72.0%) across 11 control cohorts. Pooled mean PLP intensity was 1.76 (95% CI 1.27 to 2.25) in 19 intervention cohorts and 3.45 (95% CI 2.81 to 4.08) in 11 control cohorts. For RLP, pooled prevalence was 34.7% (95% CI 26.3% to 43.1%) across 19 intervention cohorts and 62.0% (95% CI 51.4% to 72.7%) across 11 control cohorts, and pooled mean intensity was 2.05 (95% CI 1.40 to 2.70) in 21 intervention cohorts and 3.71 (95% CI 2.55 to 4.87) in 11 control cohorts. Heterogeneity was high throughout these analyses (I2 71% to 98%), and the estimates should therefore be read as descriptive summaries of the evidence base rather than as effect estimates. Forest plots for all single-arm syntheses are provided in the supplementary material.

### Sensitivity Analyses

Sensitivity analyses generally supported the direction of the primary findings. After excluding studies rated as serious overall risk of bias, PLP intensity remained lower in the intervention groups (6 studies; Hedges’ g = -0.61, 95% CI -0.78 to -0.44; p < 0.001; I2 = 0.0%), and PLP incidence remained lower (6 studies; RR = 0.57, 95% CI 0.46 to 0.71; p < 0.001; I2 = 12.5%). For RLP, exclusion of studies at serious risk of bias yielded a pooled Hedges’ g of -0.67 for intensity (6 studies; 95% CI -0.89 to -0.45; p < 0.001; I2 = 31.2%) and a pooled risk ratio of 0.52 for incidence (6 studies; 95% CI 0.36 to 0.75; p < 0.001; I2 = 60.6%).

Sensitivity analyses excluding the weakest imputations and less directly comparable outcome sources were also generated. For pain intensity these removed binary-to-mean and population-mean approximations and acute postoperative VAS scores not specific to chronic pain; for pain incidence they removed endpoints that did not use a strict no-pain definition, no-pain proportions reverse-imputed from a mean score, and studies contributing pain scores only. The direction and magnitude of effect remained consistent with the main analyses. For PLP, pooled estimates were essentially unchanged for both intensity (8 studies; Hedges’ g = -0.68, 95% CI -0.87 to -0.50; p < 0.001; I2 = 7.7%) and incidence (9 studies; RR = 0.56, 95% CI 0.48 to 0.65; p < 0.001; I2 = 0.0%). For RLP, the reduction in intensity remained substantial after excluding population-mean approximations and acute VAS values (8 studies; Hedges’ g = -0.77, 95% CI -1.04 to -0.49; p < 0.001; I2 = 48.9%), whereas RLP incidence was similar in magnitude (8 studies; RR = 0.53, 95% CI 0.41 to 0.68; p < 0.001; I2 = 43.8%). Corresponding forest plots for all sensitivity analyses are provided in the supplementary material.

### Secondary Outcomes

Secondary outcomes, comprising symptomatic neuroma and the safety outcomes, were sparsely and inconsistently reported. Meta-analysis was therefore performed only when at least two comparative studies contributed data for a given outcome. There was no clear evidence of a difference between TMR/RPNI and standard care for symptomatic neuroma (4 studies, 372 participants; RR = 0.35, 95% CI 0.11 to 1.09; p = 0.069; I2 = 24.2%), hematoma (3 studies; RR = 0.96, 95% CI 0.14 to 6.62; p = 0.97; I2 = 0.0%), infection (4 studies; RR = 0.82, 95% CI 0.45 to 1.47; p = 0.50; I2 = 0.0%), wound dehiscence (3 studies; RR = 1.83, 95% CI 0.38 to 8.87; p = 0.45; I2 = 0.0%), cardiovascular events (2 studies; RR = 0.54, 95% CI 0.07 to 4.34; p = 0.56; I2 = 0.0%), reoperation (5 studies; RR = 0.61, 95% CI 0.35 to 1.08; p = 0.091; I2 = 48.0%), or mortality (5 studies; RR = 0.63, 95% CI 0.30 to 1.32; p = 0.22; I2 = 41.7%). These estimates should be interpreted cautiously because event counts were low and confidence intervals were wide.

### Reporting Bias

Funnel plots and Egger regression tests were generated for all four main comparative pain outcomes and are provided in the supplementary material. Egger test p-values were 0.649 for PLP intensity, 0.033 for PLP incidence, 0.799 for RLP intensity, and 0.260 for RLP incidence. These results should be interpreted cautiously because each analysis included 11 studies and the evidence included was clinically heterogeneous.

### Certainty of Evidence

Using GRADE, certainty of evidence was rated as low for PLP intensity, very low for PLP incidence, very low for RLP intensity, low for RLP incidence, very low for symptomatic neuroma, and very low for safety outcomes. The main reasons for limited certainty were observational or non-randomized study designs, risk-of-bias concerns, heterogeneity for RLP intensity, possible small-study or publication bias for PLP incidence, and imprecision for symptomatic neuroma and safety outcomes. The GRADE summary of findings is provided in Supplementary Table 1.

## Discussion

### Principal Findings

This systematic review and meta-analysis suggests that prophylactic active peripheral nerve management with TMR or RPNI at the time of amputation is associated with lower post-amputation neuropathic pain compared with standard nerve management. Across comparative studies, TMR/RPNI was associated with reductions in both phantom limb pain (PLP) and residual limb pain (RLP), measured as continuous pain intensity and as binary pain incidence. The direction of effect was consistent across the four primary pain outcomes: PLP intensity, PLP incidence, RLP intensity and RLP incidence all favored TMR/RPNI.

The magnitude of association was clinically meaningful. PLP intensity was lower in the intervention groups, with a pooled standardized mean difference of approximately two thirds of a standard deviation. PLP incidence was also reduced, with intervention groups showing roughly half the risk observed in standard care groups. Similar findings were observed for RLP, although RLP intensity showed greater between-study heterogeneity.

These relative effects correspond to substantial absolute differences. Applying the pooled risk ratio to the observed control risk of 680 per 1000 gives approximately 379 per 1000 after TMR or RPNI, that is about 301 fewer patients with PLP for every 1000 treated; the corresponding figure for RLP is about 306 fewer per 1000. The supplementary single-arm syntheses point the same way, with pooled PLP prevalence of 30% in intervention cohorts against 61% in control cohorts, and pooled RLP prevalence of 35% against 62%, although those analyses do not preserve within-study comparisons and carry high heterogeneity.

The consistency of this pattern across individual studies is itself notable. Of the 44 study-level estimates contributing to the four primary analyses, 41 favored TMR or RPNI. For PLP incidence, all 11 contributing studies pointed in the same direction and between-study heterogeneity was nil.

The greater heterogeneity seen for RLP intensity is not unexpected, given variation across studies in amputation level, surgical indication, follow-up duration, pain measurement instruments, and whether reported pain scores reflected chronic RLP specifically or broader postoperative pain.

### Interpretation of Findings

The findings are biologically plausible. In standard amputation procedures, transected nerves often lack a physiological target for reinnervation; this predisposes the nerve stump to aberrant regeneration. TMR and RPNI attempt to address this mechanism directly by providing a structured target for reinnervation at the moment when the transected nerve is beginning to heal. The observed reductions in PLP and RLP are consistent with this mechanistic rationale, particularly if early reduction of peripheral ectopic input reduces later peripheral and central drivers of post-amputation pain.

Because much of the evidence included was observational, these findings should be interpreted as associations rather than causal effects. The consistency of the direction of effect supports the hypothesis that active nerve management may reduce post-amputation pain, but residual confounding and differences in patient selection cannot be excluded.

The results also suggest that the benefits of active nerve management may extend beyond symptomatic neuroma alone. PLP and RLP are complex outcomes with both peripheral and central components. By reducing peripheral nociceptive and neuropathic input early after amputation, prophylactic TMR/RPNI may reduce ongoing peripheral drivers of pain and could plausibly influence subsequent sensitization. However, the available evidence does not allow conclusions about mechanism or whether the magnitude of benefit differs across indications, amputation levels, or patient subgroups. Importantly, consistency in the direction of effect does not imply uniformity in its size. The magnitude of benefit varied appreciably across studies, and one prospectively conducted study was exactly neutral for PLP intensity.

RLP outcomes require particularly cautious interpretation. Some studies reported RLP-specific scores, whereas others reported overall pain, acute postoperative pain, or categorical pain outcomes that required conversion before pooling.

### Robustness of the Findings

The prespecified sensitivity analyses did not weaken the pooled estimates. Restricting the synthesis to studies not rated at serious overall risk of bias left every outcome essentially unchanged: Hedges’ g moved from -0.68 to -0.61 for PLP intensity and from -0.68 to -0.67 for RLP intensity, while the risk ratios moved from 0.56 to 0.57 for PLP incidence and from 0.55 to 0.52 for RLP incidence. Removing the values that depended on the weakest assumptions gave the same picture, with estimates that were unchanged for PLP and somewhat larger for RLP. This matters for interpretation: if the association were an artifact of poor methodology, it should attenuate when the weaker studies and the weaker assumptions are removed, which it did not.

This robustness speaks to internal validity, and it should not be conflated with the question of reporting bias. Excluding studies at serious risk of bias says nothing about studies that were never published, and the funnel asymmetry observed for PLP incidence is considered below. What the sensitivity analyses do establish is narrower but still useful: within the evidence that exists, the effect is not being carried by the least reliable studies or by the most aggressive imputations.

### Symptomatic Neuroma and Safety Outcomes

Symptomatic neuroma was reported with a clear numerator and denominator by only four comparative studies. The pooled estimate favored TMR/RPNI and was large in relative terms (RR 0.35, 95% CI 0.11 to 1.09), corresponding to roughly 88 fewer symptomatic neuromas per 1000 patients treated, but the confidence interval included no effect. The sparseness of this evidence is striking, because neuroma prevention is the premise of both techniques and was one of the outcomes prioritized in the protocol. The direction of effect is consistent with that premise and with the reductions observed in PLP and RLP, but confirmation will require studies that apply and report a standardized definition of symptomatic neuroma, such as Arnold’s criteria (60).

Safety outcomes were also sparsely reported. Available comparative data did not show clear evidence of differences between TMR/RPNI and standard care for hematoma, infection, wound dehiscence, cardiovascular events, reoperation, or mortality. However, confidence intervals were wide, event counts were low, and reporting was inconsistent.

This caution is particularly relevant because prophylactic TMR and RPNI can add operative steps, require specific nerve targets or muscle grafts, and may be technically demanding in complex patients. The available literature suggests that expertise, operative planning, and patient comorbidity may influence both feasibility and outcomes. Reoperation is especially difficult to interpret. Some reoperations may reflect complications of the index amputation, whereas others may represent clinically appropriate revision for pain, neuroma, wound breakdown, prosthetic fitting, or unrelated stump problems. Future studies should distinguish reoperation indications more consistently and report whether secondary procedures were performed for neuroma, pain, wound complications, bone prominence, infection, prosthetic fitting, or planned staged reconstruction.

### Clinical Implications

The results support the clinical rationale for considering TMR or RPNI as part of contemporary amputation surgery. The consistency of benefit across PLP and RLP outcomes suggests that prophylactic active nerve management at the time of amputation may offer a meaningful opportunity to reduce long-term pain burden. At the same time, TMR and RPNI procedures should not be viewed as technically interchangeable: nerve size mismatch, availability of suitable motor targets, amputation level or local tissue condition may all affect feasibility.

Implementation therefore should remain individualized. The available evidence does not yet define which patients benefit most, which amputation levels are most suitable, or whether one technique is preferable for specific scenarios.

### Methodological Considerations

This review required several outcome transformations because included studies used different instruments and reporting formats. Pain scores were standardized to a 0-10 metric, PROMIS scores were rescaled, ordinal categories were converted using midpoint assumptions, and some binary outcomes were transformed into approximate continuous values. These methods allowed a broader quantitative synthesis but necessarily introduced uncertainty. To address this, transformations were documented at study level, and sensitivity analyses were performed.

The distinction between pain prevalence, pain incidence, and absence of pain was also challenging. Some studies reported no-pain categories, while others reported mild, moderate, and severe pain, or only pain scores. In selected cases, thresholds such as no severe pain were used when another definition was unavailable. These decisions were conservative where possible, but they mean that binary outcomes should be interpreted as standardized pain presence or clinically meaningful pain burden rather than incidence.

The single-arm syntheses carry their own caveats. The proportions and mean scores across cohorts showed high heterogeneity and did not establish comparisons protecting against confounding. They are reported only to describe the observed pain burden.

### Limitations

Several limitations should be considered. First, most included studies were observational, and several were judged to be at moderate or serious risk of bias. Confounding by indication is likely: patients selected for TMR or RPNI may differ from controls in surgical indication, comorbidity or other characteristics. Second, sample sizes were often small, and some analyses included sparse event counts, leading to imprecision. Third, outcome reporting was heterogeneous. Studies differed in follow-up duration, pain instrument, pain domain definition, and whether PLP and RLP were separated. Some studies reported postoperative pain at early time points, whereas others reported longer-term chronic pain. Fourth, several of the values entering the analyses were derived rather than reported. In eight studies the standard deviation used for the standardized pain score, the no-pain proportion, or both were not available in the source publication and were carried forward as imputed values, and in a smaller number of studies continuous pain values were reconstructed from binary or categorical data. The sensitivity analyses excluding these assumptions did not change the direction or approximate magnitude of the findings, but the pooled estimates nonetheless rest in part on assumptions rather than on directly reported data. Fifth, TMR and RPNI were combined in the main intervention category. Technique-specific estimates are reported as subgroups, but only two to three studies contributed to RPNI, so the indirect comparison between techniques has low power. Sixth, safety outcomes were inconsistently reported and underpowered. Finally, the Egger test was statistically significant for PLP incidence (p = 0.033), raising the possibility of small-study effects or reporting bias for the outcome with the most precise pooled estimate.

With 11 studies this test has limited power, and the certainty of evidence for PLP incidence was nonetheless downgraded on this basis. Two features of that analysis are worth weighing alongside it: all 11 contributing studies favored TMR or RPNI, and between-study heterogeneity was nil. That pattern is more readily explained by smaller studies reporting somewhat larger effects than by a body of null studies missing from the literature, since a reservoir of unpublished null results would be expected to produce disagreement in results direction among the published studies. This interpretation cannot be tested with our data. More generally, assessment of publication bias remains limited by the small number of studies in each meta-analysis and by the observational nature of much of the evidence.

### Strengths

This review has several strengths. It was based on a prospectively registered and published protocol, used systematic methods for study selection and data extraction, and synthesized both pain intensity and pain incidence for PLP and RLP separately. The analysis retained study-level traceability of outcome transformations and incorporated sensitivity analyses to test the robustness of findings to risk of bias and weaker imputations; those analyses left the estimates essentially unchanged, which is more informative than the fact that they were performed. The near-unanimous direction of effect across contributing studies is a further strength of the underlying evidence, independent of the design limitations of individual studies. The inclusion of safety outcomes also provides a broader clinical perspective than pain efficacy alone.

### Future Research

Future studies should use standardized outcome definitions for PLP, RLP, and symptomatic neuroma. Pain outcomes should be reported separately for phantom and residual limb pain, with clear time points and validated instruments. Trials and prospective comparative cohorts should report both incidence and severity, together with standard deviations or other measures of dispersion that allow meta-analysis. Minimum clinically important differences should be prespecified when possible.

There is also a need for adequately powered comparative studies evaluating prophylactic TMR, RPNI and standard care, ideally stratified by comorbidities. Direct comparisons between TMR and RPNI are required, as current evidence does not establish the superiority of one technique or the preference in subgroups. Safety outcomes, operative time, reoperation indications, prosthetic use, functional recovery, quality of life, opioid use, and cost-effectiveness should be reported consistently.

## Conclusions

In this systematic review and meta-analysis, prophylactic TMR and RPNI performed at the time of amputation were associated with less post-amputation neuropathic pain than standard peripheral nerve management. Pain intensity was lower by approximately two thirds of a standard deviation for both phantom limb pain (PLP) and residual limb pain (RLP), and the risk of pain was roughly halved (risk ratio 0.56 for PLP and 0.55 for RLP), corresponding to about 300 fewer patients with pain for every 1000 treated.

Three features strengthen this finding. The direction of effect was near-unanimous, with 41 of the 44 study-level estimates favoring active nerve reconstruction. The pooled estimates were essentially unchanged when studies at serious risk of bias were excluded, and again when the values resting on the weakest assumptions were removed. And the effect was consistent across both pain domains and across both ways of measuring them.

Symptomatic neuroma, the mechanism on which both techniques are premised, was reported with usable data by only four comparative studies; the pooled estimate favored TMR/RPNI but was too imprecise. No study compared the two techniques directly. Compared indirectly, through their separate comparisons with standard care, they performed similarly, and no test for subgroup differences approached significance. This is an absence of evidence for a difference rather than evidence of equivalence.

Certainty in these estimates remains low to very low. That rating is driven principally by the observational design of the underlying literature, risk of bias, heterogeneous outcome definitions, and sparse safety and neuroma reporting, even though the direction of effect was consistent. The evidence supports considering TMR or RPNI at the time of amputation as a reasonable strategy in appropriate patients, while the size or distribution of the benefit awaits higher-quality studies using standardized outcomes.

## Data Availability

No data was generated by this study

## Bibliography

1. Ziegler-Graham K, MacKenzie EJ, Ephraim PL, Travison TG, Brookmeyer R. Estimating the Prevalence of Limb Loss in the United States: 2005 to 2050. Archives of Physical Medicine and Rehabilitation. 2008 Mar;89(3):422–9. doi:10.1016/j.apmr.2007.11.005

2. Alaseem A, Alanezi M, Alhuqbani MN, Aldosari ZA, Alkhunein F, Alyahya K, et al. Etiologies and Trends in Extremity Amputations: A Ten-Year Single-Center Experience. Healthcare. 2025 Sep 9;13(18):2256. doi:10.3390/healthcare13182256

3. Elaatmani M, Ahizoune A, El Maqrout A, Kharmaz M, Abouqal R, Abidi K. Anxiety and Depressive Symptoms in Moroccan Patients Following Major Lower Limb Amputation: A Three-Month Follow-Up. Cureus. 2024 May 14. doi:10.7759/cureus.60284

4. Ebrahimzadeh MH, Hariri S. Long-Term Outcomes of Unilateral Transtibial Amputations. Military Medicine. 2009 Jun;174(6):593–7. doi:10.7205/MILMED-D-02-8907

5. Huang YJ, Assi PE, Drolet BC, Al Kassis S, Bastas G, Chaker S, et al. A Systematic Review and Meta-analysis on the Incidence of Patients With Lower-Limb Amputations Who Developed Symptomatic Neuromata in the Residual Limb. Ann Plast Surg. 2022 May;88(5):574–80. doi:10.1097/SAP.0000000000002946

6. Wu H, Saini C, Medina R, Hsieh SL, Meshkati A, Sung K. Pain without presence: a narrative review of the pathophysiological landscape of phantom limb pain. Front Pain Res. 2025 Feb 18;6:1419762. doi:10.3389/fpain.2025.1419762

7. Janes LE, Fracol ME, Dumanian GA, Ko JH. Targeted Muscle Reinnervation for the Treatment of Neuroma. Hand Clinics. 2021 Aug;37(3):345–59. doi:10.1016/j.hcl.2021.05.002

8. Markewych AN, Suvar T, Swanson MA, Graca MJ, Lubenow TR, McCarthy RJ, et al. Approaches to neuropathic amputation-related pain: narrative review of surgical, interventional, and medical treatments. Reg Anesth Pain Med. 2024 Dec;49(12):889–99. doi:10.1136/rapm-2023-105089

9. Pet MA, Ko JH, Friedly JL, Smith DG. Traction Neurectomy for Treatment of Painful Residual Limb Neuroma in Lower Extremity Amputees. Journal of Orthopaedic Trauma. 2015 Sep;29(9):e321–5. doi:10.1097/BOT.0000000000000337

10. Richards JT, Baird MD, Tintle SM, Souza JM, Renninger CH, Potter BK. Peripheral Nerve Management in Extremity Amputations. Orthopedic Clinics of North America. 2022 Apr;53(2):155–66. doi:10.1016/j.ocl.2022.01.002

11. Campo-Prieto P, Rodríguez-Fuentes G. Effectiveness of mirror therapy in phantom limb pain: a literature review. Neurología (English Edition). 2022 Oct;37(8):668–81. doi:10.1016/j.nrleng.2018.08.005

12. Biloshytsky VV, Biloshytska MV. Pharmacological and interventional treatment of phantom pain. Ukr Neurosurg J. 2025 Mar 31;31(1):3–11. doi:10.25305/unj.318305

13. Zimbulis AJ, An VVG, Symes M, Duraku LS, Gaston RG, Eberlin KR, et al. Targeted Muscle Reinnervation Compared to Standard Peripheral Nerve Management Following Amputation: A Systematic Review and Meta-Analysis. Hand (New York, NY). 2024 Oct 29;15589447241284811. doi:10.1177/15589447241284811

14. Hooper RC, Cederna PS, Brown DL, Haase SC, Waljee JF, Egeland BM, et al. Regenerative Peripheral Nerve Interfaces for the Management of Symptomatic Hand and Digital Neuromas. Plastic and Reconstructive Surgery - Global Open. 2020 Jun;8(6):e2792. doi:10.1097/GOX.0000000000002792

15. Kubiak CA, Kemp SWP, Cederna PS. Regenerative Peripheral Nerve Interface for Management of Postamputation Neuroma. JAMA Surg. 2018 Jul 1;153(7):681. doi:10.1001/jamasurg.2018.0864

16. Roubaud MJ, Babu A, Moon BS, Lewis VO. Targeted Muscle Reinnervation and Regenerative Peripheral Nerve Interface: The Evolving Landscape in the Treatment of Postamputation Pain and Prosthetics. J Am Acad Orthop Surg. 2026 Feb 1;34(3):e324–34. doi:10.5435/JAAOS-D-25-00252

17. Savitz BL, Dean YE, Popa NK, Cornely RM, Byers V, Gutama BW, et al. Targeted Muscle Reinnervation and Regenerative Peripheral Nerve Interface for Myoelectric Prosthesis Control: The State of Evidence. Ann Plast Surg. 2025 Jun;94(6S):S572–6. doi:10.1097/SAP.0000000000004273

18. Maile N, Ferrante L, Farina D, Al Dabbass U, Aszmann OC, Tereshenko V. Targeted Muscle Reinnervation for Intuitive Prosthetic Control. Muscle and Nerve. 2026 Jul 7;mus.70340. doi:10.1002/mus.70340

19. Raasveld FV, Mayrhofer-Schmid M, Johnston BR, Harhaus-Waehner L, Zuidam JM, Valerio IL, et al. Targeted Muscle Reinnervation for Management and Prevention of Symptomatic Neuroma. Muscle and Nerve. 2026 Jul 4;mus.70335. doi:10.1002/mus.70335

20. Cathey JM, Klifto KM, Anderson JL, Li SY, Saltzman EB, Li NY. Targeted Muscle Reinnervation at the Time of Major Limb Amputation Reduces Long-term Use and Dependence for Opioid Analgesic Therapy: A Multicenter Propensity-matched Study. Annals of Surgery. 2026 Aug;284(2):436–44. doi:10.1097/SLA.0000000000006697

21. De Lange JWD, Hundepool CA, Power DM, Rajaratnam V, Duraku LS, Zuidam JM. Prevention is better than cure: Surgical methods for neuropathic pain prevention following amputation– A systematic review. Journal of Plastic, Reconstructive & Aesthetic Surgery. 2022 Mar;75(3):948–59. doi:10.1016/j.bjps.2021.11.076

22. Mauch JT, Kao DS, Friedly JL, Liu Y. Targeted muscle reinnervation and regenerative peripheral nerve interfaces for pain prophylaxis and treatment: A systematic review. PMCR. 2023 Nov;15(11):1457–65. doi:10.1002/pmrj.12972

23. Yuan M, Gallo M, Gallo L, Huynh MH, McRae M, McRae MC, et al. Targeted Muscle Reinnervation and Regenerative Peripheral Nerve Interfaces Versus Standard Management in the Treatment of Limb Amputation: A Systematic Review and Meta-Analysis. Plast Surg (Oakv). 2024 May;32(2):253–64. doi:10.1177/22925503221107462

24. Higgins JPT, Lasserson T, Chandler J, Tovey D, Thomas J, Flemyng E, Churchill R. Methodological Expectations of Cochrane Intervention Reviews. Cochrane: London, Version February 2021. https://community.cochrane.org/mecir-manual.

25. Page MJ, Moher D, Bossuyt PM, Boutron I, Hoffmann TC, Mulrow CD, et al. PRISMA 2020 explanation and elaboration: updated guidance and exemplars for reporting systematic reviews. BMJ. 2021 Mar 29;n160. doi:10.1136/bmj.n160

26. Page MJ, McKenzie JE, Bossuyt PM, Boutron I, Hoffmann TC, Mulrow CD, et al. The PRISMA 2020 statement: an updated guideline for reporting systematic reviews. BMJ. 2021 Mar 29;n71. doi:10.1136/bmj.n71

27. Del Moral Preciado J, Gurpegui D, Royo M, Hontanilla B. Targeted Muscle Reinnervation (TMR) or Regenerative Peripheral Nerve Interface (RPNI) for pain prevention in patients with limb amputation: A protocol for a systematic review and meta-analysis. PLoS One. 2026 Jun 5;21(6):e0350845. doi:10.1371/journal.pone.0350845

28. Sterne JAC, Savović J, Page MJ, Elbers RG, Blencowe NS, Boutron I, et al. RoB 2: a revised tool for assessing risk of bias in randomised trials. BMJ. 2019 Aug 28;l4898. doi:10.1136/bmj.l4898

29. Sterne JA, Hernán MA, Reeves BC, Savović J, Berkman ND, Viswanathan M, et al. ROBINS-I: a tool for assessing risk of bias in non-randomised studies of interventions. BMJ. 2016 Oct 12;i4919. doi:10.1136/bmj.i4919

30. Alexander JH, Jordan SW, West JM, Compston A, Fugitt J, Bowen JB, et al. Targeted muscle reinnervation in oncologic amputees: Early experience of a novel institutional protocol. Journal of Surgical Oncology. 2019 Sep;120(3):348–58. doi:10.1002/jso.25586

31. Frantz TL, Everhart JS, West JM, Ly TV, Phieffer LS, Valerio IL. Targeted Muscle Reinnervation at the Time of Major Limb Amputation in Traumatic Amputees: Early Experience of an Effective Treatment Strategy to Improve Pain. JBJS Open Access. 2020;5(2):e0067. doi:10.2106/JBJS.OA.19.00067

32. T Kumar A, Raajeshwaran MA, Bagaria DK, Mir N, Sagar S, Alam J, et al. Effectiveness of pre-emptive targeted muscle reinnervation on post-amputation pain in patients undergoing above knee amputation: A randomized controlled trial. J Trauma Acute Care Surg. 2026 Jan 30. doi: 10.1097/TA.0000000000004907.

33. Raasveld FV, Mayrhofer-Schmid M, Johnston BR, Gomez-Eslava B, Hoftiezer YAJ, Liu WC, et al. Targeted muscle reinnervation at the time of amputation to prevent the development of neuropathic pain. Journal of Plastic, Reconstructive & Aesthetic Surgery. 2024 Oct;97:13– 22. doi:10.1016/j.bjps.2024.07.055

34. Reid RT, Johnson CC, Gaston RG, Loeffler BJ. Impact of Timing of Targeted Muscle Reinnervation on Pain and Opioid Intake Following Major Limb Amputation. Hand (New York, NY). 2024 Mar;19(2):200–5. doi:10.1177/15589447221107696

35. Smith TP, Cognetti DJ, Cook A, Lynch TB, Alderete JF, Lybeck DO, et al. Similar rates of reoperation for neuroma after transtibial amputations with and without targeted muscle reinnervation. OTA International. 2024 Mar;7(1). doi:10.1097/OI9.0000000000000297

36. Elmorsi R, Shin A, Adelman DM, Mericli AF, Roubaud MJ. 52. Efficacy of Targeted Muscle Reinnervation and Regenerative Peripheral Nerve Interface in Reducing Pain in Cancer Amputees: A Patient-reported Outcomes Study. Plastic and Reconstructive Surgery - Global Open. 2025 May;13(S2):26. doi:10.1097/01.GOX.0001114052.82447.3e

37. Mathieu L, Redais C, Diner C, Lemaire-Petit A, Milaire A, Chataigneau A, et al. Curative and preemptive treatment of amputee pain by targeted muscle reinnervation: experience from a French military trauma center. Eur J Trauma Emerg Surg. 2025 Dec;51(1):37. doi:10.1007/s00068-024-02701-w

38. StataCorp. 2019. Stata: Release 16. Statistical Software. College Station, TX: StataCorp LLC.

39. DerSimonian R, Laird N. Meta-analysis in clinical trials revisited. Contemporary Clinical Trials. 2015 Nov;45:139–45. doi:10.1016/j.cct.2015.09.002

40. Obinero CG, Green JC, Swiekatowski KR, Obinero CV, Manisundaram A, Greives MR, et al. Surgical Complications after Targeted Muscle Reinnervation at a Safety-Net Hospital. J Reconstr Microsurg. 2025 Jul;41(06):531–9. doi:10.1055/a-2435-7410

41. Mahmoud MA, Toyoda Y, McGraw JR, Sulkar R, Saikali L, Lopez S, et al. 40. Development Of Neuropathic Pain After Primary TMR And RPNI In Lower Extremity Amputation: A 4-year Retrospective Analysis Of Prognostic Factors. Plastic and Reconstructive Surgery - Global Open. 2024 May;12(S5):25. doi:10.1097/01.GOX.0001018176.36476.78

42. Berberoglu I, Kemp SWP, Hooper RC. Regenerative Peripheral Nerve Interface (RPNI) for the Management of Postamputation Pain in Oncology Patients. Plastic & Reconstructive Surgery. 2025 Jul 15. doi:10.1097/PRS.0000000000012312

43. Kubiak CA, Kemp SWP, Cederna PS, Kung TA. Prophylactic Regenerative Peripheral Nerve Interfaces to Prevent Postamputation Pain. Plastic & Reconstructive Surgery. 2019 Sep;144(3):421e–30e. doi:10.1097/PRS.0000000000005922

44. Chang BL, Mondshine J, Attinger CE, Kleiber GM. Targeted Muscle Reinnervation Improves Pain and Ambulation Outcomes in Highly Comorbid Amputees. Plastic & Reconstructive Surgery. 2021 Aug;148(2):376–86. doi:10.1097/PRS.0000000000008153

45. Chang BL, Hill AL, Mondshine J, Harbour PW, Episalla NC, Attinger CE, et al. Primary Targeted Muscle Reinnervation in Above-Knee Amputations in Patients with Unsalvageable Limbs from Limb-Threatening Ischemia or Infection. J Reconstr Microsurg. 2024 Feb;40(02):109–17. doi:10.1055/a-2086-0395

46. Pejkova S, Nikolovska B, Srbov B, Tusheva S, Jovanoski T, Jovanovska K, et al. Prophylactic Regenerative Peripheral Nerve Interfaces in Elective Lower Limb Amputations. PRILOZI. 2022 Mar 1;43(1):41–8. doi:10.2478/prilozi-2022-0004

47. Guyatt GH, Oxman AD, Vist GE, et al. GRADE: an emerging consensus on rating quality of evidence and strength of recommendations. BMJ. 2008;336(7650):S24–6)

48. O’Brien AL, Jordan SW, West JM, Mioton LM, Dumanian GA, Valerio IL. Targeted Muscle Reinnervation at the Time of Upper-Extremity Amputation for the Treatment of Pain Severity and Symptoms. The Journal of Hand Surgery. 2021 Jan;46(1):72.e1-72.e10. doi:10.1016/j.jhsa.2020.08.014

49. Huffman A, Schneeberger S, Goodyear E, West JM, O’Brien AL, Scharschmidt TJ, et al. Evaluating hip disarticulation outcomes in a 51-patient series. Journal of Orthopaedics. 2022 May;31:117–20. doi:10.1016/j.jor.2022.04.008

50. Shammas RL, Azoury SC, Sergesketter AR, Lee HJ, Poehlein E, Othman SE, et al. Primary Targeted Muscle Reinnervation after Below-Knee Amputation Is Not Associated with an Increased Risk of Major or Minor Surgical Complications: A Multi-Institutional, Propensity Score–Matched Analysis. Plastic & Reconstructive Surgery. 2022 Sep;150(3):589–98. doi:10.1097/PRS.0000000000009441

51. Vaeth AM, Kochheiser M, Qin N, Wei L, Truong AY, Hoellwarth JS, et al. D123. Advances in Neuroma Prevention: A Single Institutional Experience With TMR And TMRPNI in Lower Limb Amputees. Plastic and Reconstructive Surgery - Global Open. 2025 May;13(S2):143. doi:10.1097/01.GOX.0001114984.51049.58

52. Lu V, Zhou A, Krkovic M. Variation on a technique for the intra-muscular insertion of nerve endings to minimise neuropathic and residual pain in lower limb amputees: a retrospective cohort study. Eur J Orthop Surg Traumatol. 2022 May 25;33(4):1299–306. doi:10.1007/s00590-022-03281-4

53. Valerio IL, Dumanian GA, Jordan SW, Mioton LM, Bowen BJ, West JM, et al. Preemptive Treatment of Phantom and Residual Limb Pain with Targeted Muscle Reinnervation at the Time of Major Limb Amputation. Journal of the American College of Surgeons. 2019 Mar;228(3):217–26. doi:10.1016/j.jamcollsurg.2018.12.015

54. Li AT, Garcia N, Angliss M, Paul E, Gray S, Bruscino-Raiola F. Acute versus non-acute targeted muscle reinnervation for pain control following major limb amputation: A comparative study. Journal of Plastic, Reconstructive & Aesthetic Surgery. 2024 Jul;94:229– 37. doi:10.1016/j.bjps.2024.05.011

55. Pet MA, Ko JH, Friedly JL, Mourad PD, Smith DG. Does Targeted Nerve Implantation Reduce Neuroma Pain in Amputees? Clinical Orthopaedics & Related Research. 2014 Oct;472(10):2991–3001. doi:10.1007/s11999-014-3602-1

56. Raasveld FV, Hoftiezer YAJ, Gomez-Eslava B, McCarty J, Valerio IL, Heng M, et al. Early Postoperative Pain Course following Primary and Secondary Targeted Muscle Reinnervation: A Temporal Description of Pain Outcomes. J Reconstr Microsurg. 2025 Jul;41(06):459–68. doi:10.1055/a-2404-7732

57. Goodyear EG, O’Brien AL, West JM, Huayllani MT, Huffman AC, Souza JM, et al. Targeted Muscle Reinnervation at the Time of Amputation Decreases Recurrent Symptomatic Neuroma Formation. Plastic & Reconstructive Surgery. 2024 Jan;153(1):154–63. doi:10.1097/PRS.0000000000010692

58. Bowen JB, Ruter D, Wee C, West J, Valerio IL. Targeted Muscle Reinnervation Technique in Below-Knee Amputation. Plastic & Reconstructive Surgery. 2019 Jan;143(1):309–12. doi:10.1097/PRS.0000000000005133

59. O’Brien AL, West JM, Gokun Y, Janse S, Schulz SA, Valerio IL, et al. Longitudinal Durability of Patient-Reported Pain Outcomes after Targeted Muscle Reinnervation at the Time of Major Limb Amputation. Journal of the American College of Surgeons. 2022 May;234(5):883–9. doi:10.1097/XCS.0000000000000117

60. Arnold DMJ, Wilkens SC, Coert JH, Chen NC, Ducic I, Eberlin KR. Diagnostic Criteria for Symptomatic Neuroma. Ann Plast Surg. 2019 Apr;82(4):420–7. doi:10.1097/SAP.0000000000001796

